# Estimation of effects of contact tracing and mask adoption on COVID-19 transmission in San Francisco: a modeling study

**DOI:** 10.1101/2020.06.09.20125831

**Authors:** Lee Worden, Rae Wannier, Seth Blumberg, Alex Y. Ge, George W. Rutherford, Travis C. Porco

## Abstract

The current COVID-19 pandemic has spurred concern about what interventions may be effective at reducing transmission. The city and county of San Francisco imposed a shelter-in-place order in March 2020, followed by use of a contact tracing program and a policy requiring use of cloth face masks. We used statistical estimation and simulation to estimate the effectiveness of these interventions in San Francisco. We estimated that self-isolation and other practices beginning at the time of San Francisco’s shelter-in-place order reduced the effective reproduction number of COVID-19 by 35.4% (95% CI, −20.1%–81.4%). We estimated the effect of contact tracing on the effective reproduction number to be a reduction of approximately 44% times the fraction of cases that are detected, which may be modest if the detection rate is low. We estimated the impact of cloth mask adoption on reproduction number to be approximately 8.6%, and note that the benefit of mask adoption may be substantially greater for essential workers and other vulnerable populations, residents return to circulating outside the home more often. We estimated the effect of those interventions on incidence by simulating counterfactual scenarios in which contact tracing was not adopted, cloth masks were not adopted, and neither contact tracing nor cloth masks was adopted, and found increases in case counts that were modest, but relatively larger than the effects on reproduction numbers. These estimates and model results suggest that testing coverage and timing of testing and contact tracing may be important, and that modest effects on reproduction numbers can nonetheless cause substantial effects on case counts over time.

## Introduction

The current COVID-19 pandemic has spurred concern about what interventions may be effective at reducing transmission. In San Francisco and several nearby counties, an official order to shelter in place^1^ took effect on March 17, 2020, directing people to stay at home and avoid all but essential gatherings, travel, and business. Schools and some day care facilities and public places such as parks have additionally been closed. On March 19, a similar stay-at-home order was issued covering the entire state of California.^2^ More recently, San Francisco has introduced a program of contact tracing, and has begun to require the use of cloth face masks beginning April 18, 2020.^3^ At the time of this writing, the shelter-in-place order has been extended through the month of May. The accumulation of new COVID-19 cases in San Francisco appears to have slowed since late March.

Because of the danger of widespread transmission of SARS-Cov-2 in terms of illness and loss of life and potential overloading of health care resources, and hardships associated with self-isolation, it can be important to estimate the effect of epidemic control measures on transmission of the disease.

We used simulation models to estimate the effect of the March 17 shelter-in-place and related changes on COVID-19 transmission rates in San Francisco.

While other models have attempted to estimate the impact of quarantine and isolation [5–8, 10, 18–21, 23, 25, 27, 30–38, 40, 44, 46–48] or used complex models to look at the impact of various interventions [11–13, 39, 45] none have yet combined mask adoption and test-and-treat fit to local geographic data. We estimated the effects of contact tracing and widespread mask adoption on COVID-19 transmission rates in San Francisco, and projected case counts forward four weeks with and without contact tracing and mask adoption to estimate their effect on numbers of cases.

## Methods

### Data

County-level cumulative case counts are reported by individual counties, frequently on a daily basis. These have been aggregated by the New York Times.^4^

### Transmission model

Estimation of parameters and projection of case counts into the future were done using a stochastic renewal process model that simulates a population of infected individuals, some of whom are detected and included in reported case counts. The key assumptions were as follows.

#### The time-varying reproduction number (*R*_*t*_) is unknown

The model assumed a constant value of *R*_*t*_ ranging from 0.3 to 5 until March 17, 2020, and three separate constant values of *R*_*t*_ in the same range after March 17, with discontinuous changes on April 8 and April 18, reflecting the adoption of San Francisco’s shelter in place, contact tracing, and cloth mask policies. It produced estimates of *R*_*t*_ by selecting from these possibilities whichever values were best able to match the case counts reported to date in the Bay Area.

#### The incubation period is approximately 5–6 days

The incubation interval was modeled using a log-normal distribution with mean 5.6 days and standard deviation 4.2 days.

#### Transmission can begin before the onset of symptoms

Transmission was assumed to occur roughly from two days before symptom onset to several days after system onset.

#### Delays in case reporting are variable

Reporting of cases was assumed to occur after a variable period of delay, most often around 4–5 days after symptom onset, but with a tail extending out to several weeks after symptom onset.

#### The rate of reporting of true COVID-19 cases is unknown

The reporting rate was allowed to range from 1.1% reporting (the lower bound value of 1 in 85 which appeared in the initial draft of a recent Santa Clara county study [3], dated 17 April 2020, and subsequently revised) to 60% reporting. The model selects whichever values are best able to reproduce the case counts reported to date in the Bay Area.

For more detail, see the Appendix.

### Estimation of effect of contact tracing on transmission

We estimated the impact of contact tracing on transmission using the same estimates of the incubation period, timing of transmission events, and delay from symptom onset to case detection as in the above model (see Appendix for details).

Contact tracing consists of identifying contacts *y* who may have been infected by a confirmed case *x* and attempting to isolate them to prevent them from transmitting to other people. If contact tracing happens the day a case *x* is detected, the success of contact tracing can be modeled by estimating how often the detection date of case *x* predates the dates of transmission from contacts *y*.

The interval from symptom onset of *x* to dates of transmission from contacts *y* is a sum of random variables (Figure 1): the time from symptom onset of *x* to transmission to a contact *y* (which may be negative), plus the incubation interval to symptom onset of *y*, plus the interval from that to transmission from *y*.

**Figure 1:**
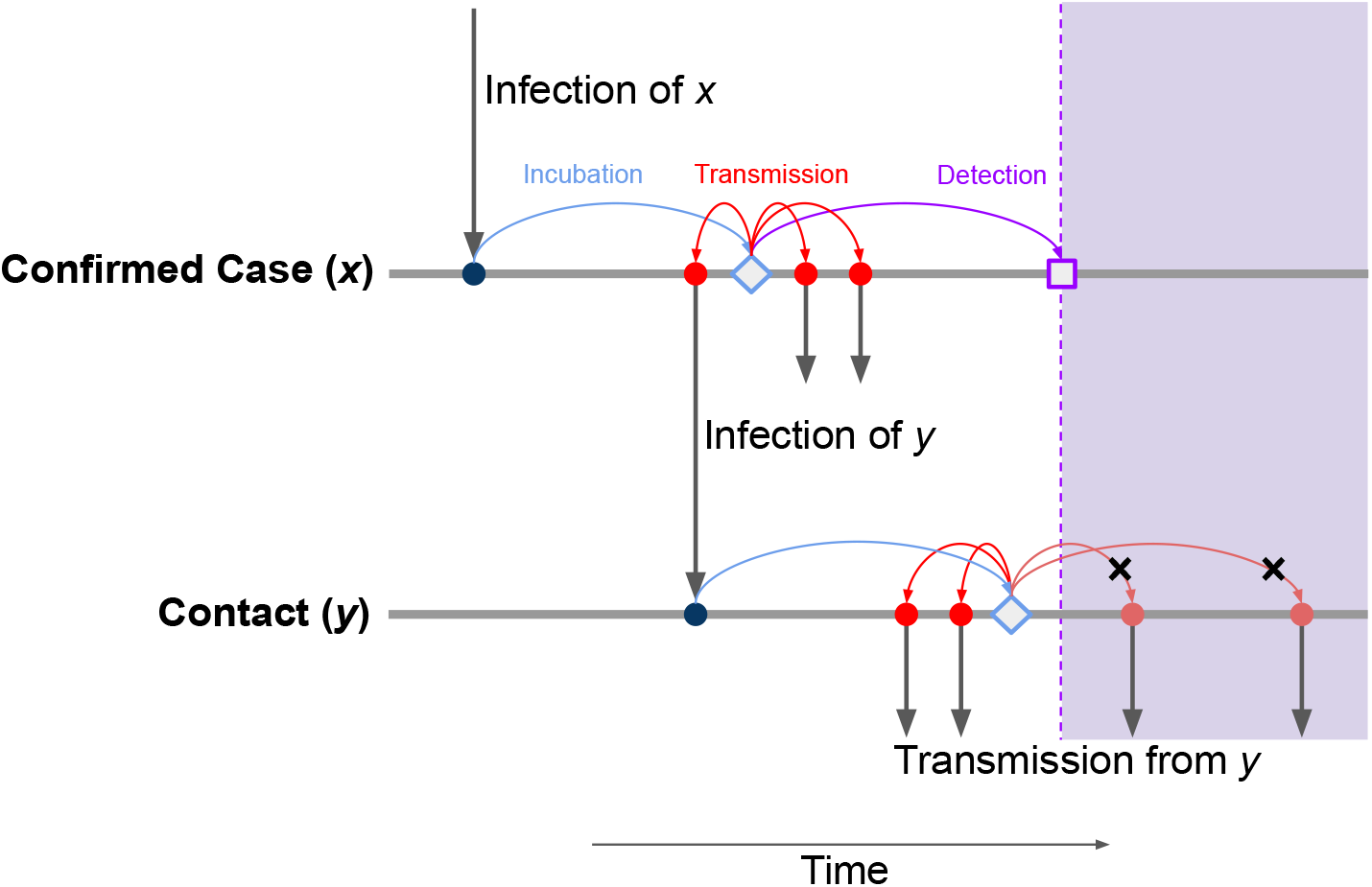
Schematic of timing of events involved in contact tracing. Contact tracing of a confirmed case (*x*) aims at preventing transmission from contacts (*y*) of that case. It can succeed when potential transmission events occur after case detection (lavender shaded area). This requires the interval from symptom onset to case confirmation (purple arrow) to be shorter than the time to transmission to a contact (red arrows, upper line) plus the incubation period of a contact (light blue arrow, lower line) plus the time to transmission from a contact (red arrows, lower line).

By Monte Carlo sampling we found that the detection interval is exceeded by the interval to transmission from contacts in approximately 44% of cases.

We conducted sensitivity analysis of these assumptions by altering the mean time to case detection downward from 8.9 days to 5 days, and upward to 13 days, while leaving the logSD parameter unchanged. We found detection predating transmission to contacts of contacts in 64% and 29% of cases, respectively.

Thus, we assumed that contact tracing can prevent 44% of transmission from a primary case. We modeled contact tracing by a reduction in reproduction number of 0.44 *p*_*d*_, where *p*_*d*_ is the probability that a given case is detected.

San Francisco’s contact tracing program, aside from a brief early rollout, went into effect gradually in early April 2020. We approximate this for modeling purposes by a start date of April 8, 2020. We used the above estimate to model what would have happened in the absence of the contact tracing program by modeling case counts assuming that reproduction numbers are inflated by a factor of 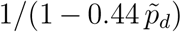 from April 8 onward, where 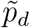 is the rate of case detection estimated by the model.

### Estimation of the effect of cloth mask adoption on transmission

We estimated the effect of wearing cloth facial masks in the community and workplaces in reducing transmission [2, 15, 17, 26, 29, 41]. We assumed the population to be 80% compliant with mask wearing policy, that cloth masks reduce the probability of an infected individual transmitting to an uninfected and unmasked individual by 30%, and that an uninfected individual is only 10% protected by wearing a cloth mask, based on studies showing that the primary benefit of wearing cloth or surgical masks is to protect the environment from the wearer rather than the wearer from the environment [2, 15, 41]. These probabilities were combined to yield an overall reduction of transmission probability in workplaces as well as the community of 30.1%.

These estimates were then used to estimate the overall population reduction in transmission when mask wearing is implemented in the communities and workplaces. We used estimates of contact and transmission rates from influenza models to estimate the contribution of the neighborhood and workplace to effective transmission [14]. We also used the mean length of the infectious period and a mean household size of 2.69 in the San Francisco Bay Area [1] to estimate the reduction in effective transmission within the household due to depletion of susceptibles over the infectious period, to more accurately estimate effective contribution of household transmission to the overall transmission. Based upon a 75% reduction [12] in effective contact rates in the workplace and community domains under Shelter-in-Place orders, we estimate that 71% of transmission occurs in households, 17% in the neighborhood and 12% in the workplace. From extending these assumption, we estimate that masks will be worn during only 29% of otherwise effective transmissions, and thus masks will reduce effective transmission rate by 8.6%.

We note that this estimate reflects assumptions that the interactions affected by mask usage—contacts in neighborhoods and workplaces—are substantially reduced by the practice of sheltering in place. If communities return to work and to circulating in public more, the use of masks will surely have more effect. We note also that the benefit of masks may be much greater among people who are less able to self isolate, such as essential workers or unhoused individuals.

We used this estimate to model the effect of mask use on epidemic dynamics by modeling a scenario in which masks were not adopted on April 18, the start date of San Francisco’s cloth mask policy, by assuming that reproduction numbers are inflated from April 18 on, by a factor of 1*/*(1 − 0.086), or approximately 1.094, and by modeling the case in which both contact tracing and cloth masks were not adopted, by combining this inflation with the inflation described above for contact tracing.

## Results

We estimated effective instantaneous reproduction numbers *R*_*t*_ under the above mentioned model assumptions of piecewise constant values, reflecting a possible changing effect of social distancing and other protective measures (Figure 2). Our model estimated a decline in reproduction numbers, from a mean value of 1.87 before 17 March (95% central interval, 1.38–2.62) to 1.15 (95% central interval, 0.37–1.86) between 17 March and 7 April. San Francisco’s shelter-in-place policy and related measures were estimated to have led to a mean 35.4% reduction in effective reproduction number (95% central interval, −20.1%–81.4%).

**Figure 2:**
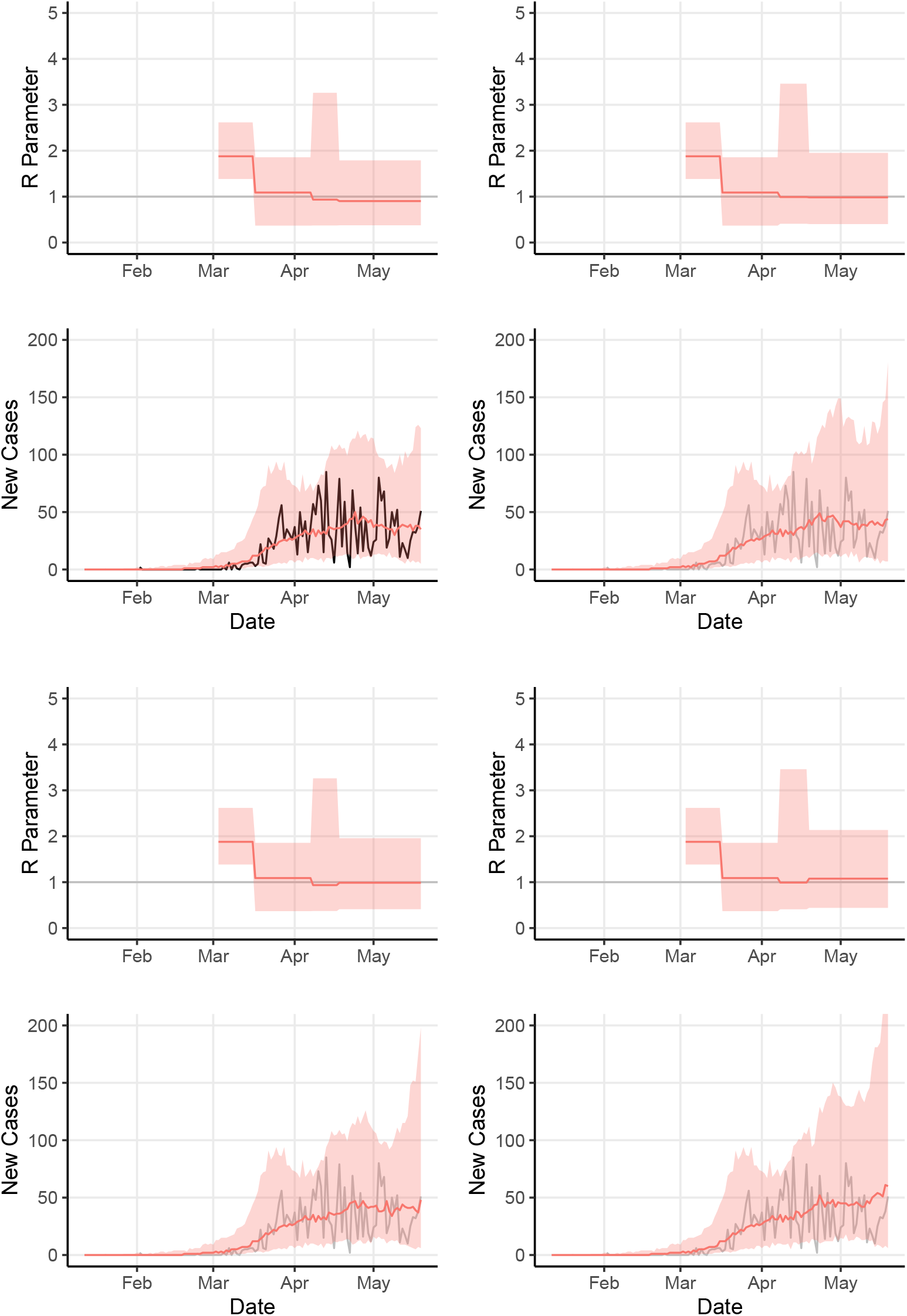
Model estimates of effective *R*_*t*_ and number of cases, fit to San Francisco counts and in three counterfactual scenarios. Top Left: model fit to San Francisco counts; Top Right: supposing contact tracing did not begin April 8, 2020; Bottom Left: supposing adoption of cloth masks did not begin April 18, 2020; Bottom Right: supposing both contact tracing and cloth mask adoption did not happen. Model assumes *R*_*t*_ constant except for instantaneous changes on March 17, April 8, and April 18, 2020. Ribbons show mean and 95% central interval of effective *R*_*t*_ values and median and 95% central interval of model case counts, weighted by importance sampling (see Methods).

Second, we estimated how the recent course of the outbreak might have been altered by the adoption of contact tracing and masks. We used the renewal process model to generate estimates of time-varying reproduction numbers, overall incidence, and other parameters from daily reported case counts from January 2020 through May 19, 2020. We compared the model fit to true reported cases to three counterfactual scenarios for Bay Area transmission during those dates (Figures 2 and 3). The model’s mean estimated reproduction number *R*_*t*_ declined from 1.85 to 1.13 in San Francisco following widespread sheltering in place, consistent with substantial reduction in transmission as a direct result of the intervention. A median count of 36.7 reported cases per day was estimated over the final week May 13–May 19, 2020 in the fit to San Francisco reported data, compared to medians of 40.9 daily cases in the counterfactual scenario without contact tracing, 41.9 daily cases in the scenario without mask adoption, and 52.7 daily cases in the scenario without both contact tracing and masks.

**Figure 3:**
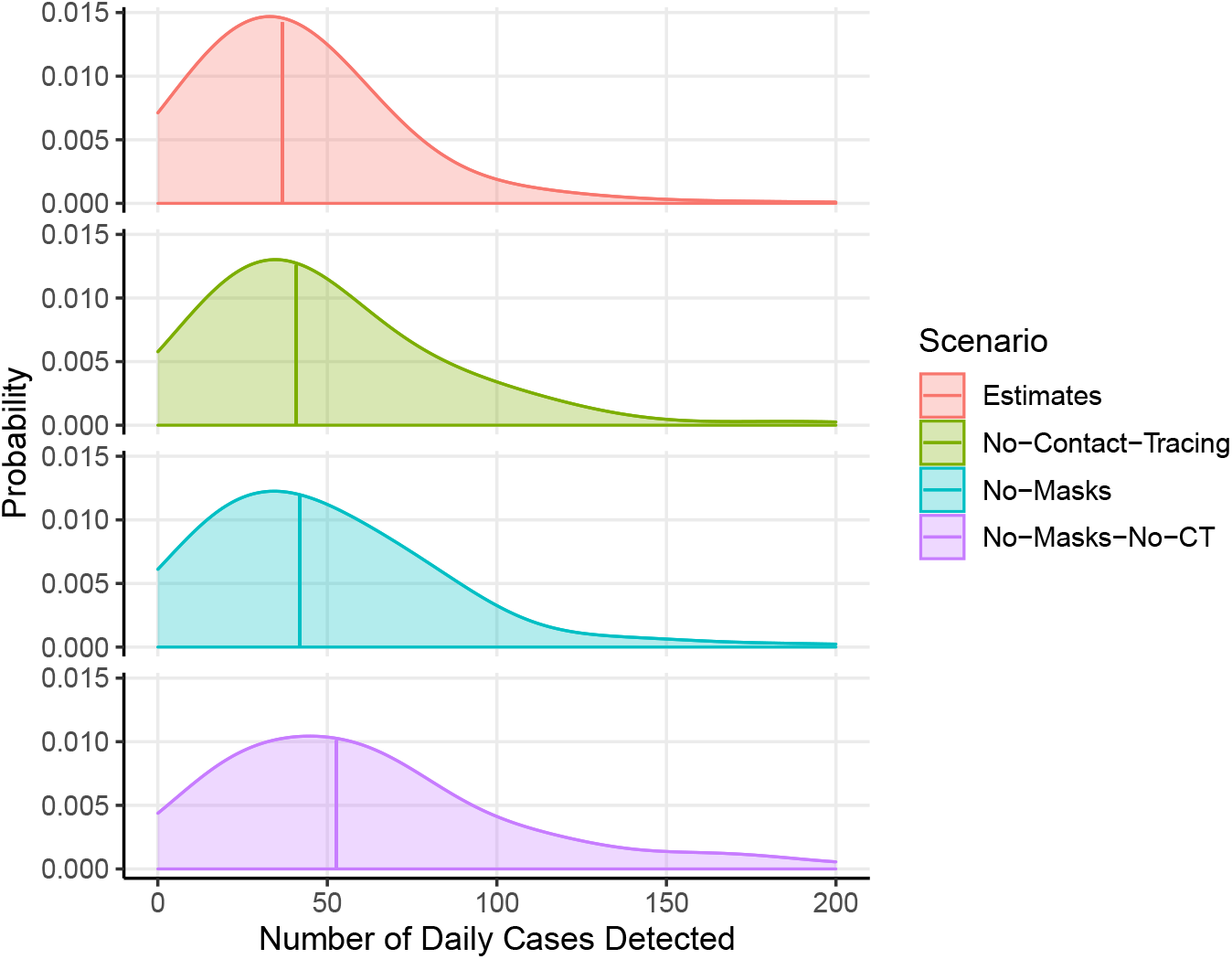
Distributions of model case counts over the week May 13–May 19, 2020, with and without interventions. Estimated from model outcomes using Gaussian kernel smoothing with bandwidth of 20 cases. Vertical bars represent median number of cases.

## Discussion

Despite a substantial estimated decline in transmission rate, represented by change in reproduction number at the time of sheltering in place, San Francisco has not achieved unambiguous subcriticality and thus we cannot conclude that the epidemic is ebbing.

We have estimated that reductions in transmission associated with both the contact tracing program and the adoption of cloth masks may be modest, but we note that because the reproduction number *R*_*t*_ is an exponential growth rate, modest differences in *R*_*t*_ can lead to substantial differences in the course of an outbreak.

We also note that while the effects of both interventions on *R*_*t*_ may be modest, it is for different reasons. The impact of contact tracing on the overall rate of transmission may be smaller than it could be for two reasons: because contact tracing is carried out on individuals who receive positive test results, who may be a fraction of the true number of cases, and because fast spread of the infection combined with delays in case detection may make it difficult to isolate contacts before they pass on the virus.

Cloth masks, on the other hand, which can reduce transmission substantially in neighborhood and workplace settings, are estimated to have a modest impact on transmission during shelter in place measures simply because neighborhood and workplace contacts are contributing less transmission. It is likely that masks can reduce transmission more substantially if social distancing practices are lessened, and that they can be important in reducing transmission to essential workers, including healthcare workers, and among populations who may have substantial contact with strangers such as unhoused communities.

Our estimate for contact tracing points to the importance of widespread testing in controlling the spread of this disease. In addition, it highlights the importance of timing to the effectiveness of contact tracing. Contact tracing may be most effective in interrupting transmission when applied to seed cases who are identified very soon after symptom onset (if not before), and to their most recent contacts. There might be some benefit to contact tracing some individuals prospectively without waiting for a positive test result, to gain the advantage of time at the cost of some accuracy. We note also that contact tracing has multiple goals and benefits beyond reducing transmission, including locating cases and giving people access to guidance and testing and treatment.

We note that our group found a similar modest effect of contact tracing in measles control, because the much higher transmissibility means that much transmission may be to casual or anonymous contacts who are hard to identify, as well as because high vaccination rates imply that many contacts will have already been vaccinated [24].

This model exhibits the following limitations. We assumed constant reproduction numbers *R*_*t*_ before and after the adoption of San Francisco’s shelter in place policy, so this model cannot detect more detailed changes in *R*_*t*_. The model assumes uniform dynamics across all cases in a simulation of a county, not accounting for specific geographic and demographic heterogeneity. The city and county of San Francisco is modeled in isolation, without migration or transmission across county lines (since we anticipate more transmission within than between counties). Detection rates are assumed to be constant per capita; a gradually rising detection rate can cause an upward bias [28]. If testing is limited by available resources, the assumption of a per capita rate may not be justified. Alternative hypotheses for the unknown interval distributions of generation time and time to detection should be evaluated. In particular, the impact of the amount of transmission occurring before onset of symptoms, and the tail of long times to detection, should be examined carefully. The distribution of generation intervals is affected by censoring of transmission after cases’ detection dates, which shifts generation intervals to shorter times, so that our baseline interval distribution should ideally be for generation times in the absence of detection, but our estimates are taken from populations with some amount of detection. This could lead to bias toward shorter generation times than is accurate. Heterogeneity in transmission rates between communities within the city, and across racial differences, appears to be very important as vulnerable populations and specifically people of color are disproportionately affected, and should be considered explicitly where possible.

## Data Availability

This manuscript used only publicly available data.

## Acknowledgements

LW and TCP acknowledge support from the US NIH NIGMS MIDAS program, R01-GM130900. The views, opinions, assumptions and conclusions or any other information set out in this article are solely those of the authors and do not necessarily reflect those of the NIH.

## Appendix

### Stochastic model implementation

The simulation model constructed a stochastic time series of case counts by day, based on assumptions about transmission rates and time intervals. Let the number of transmission events (equivalently, the number of cases infected) on day *i* be denoted *t*_*i*_. Each run of the simulation began with some number of index cases infected on day 1. For each day *i*, the model estimated secondary cases from the *t*_*i*_ cases infected on day *i*. Each secondary case was assigned an infection date *j > i*, and it was added to the count *t*_*j*_.

Case detection was modeled as follows. Let *d*_*i*_ denote the number of cases that would be detected on day *i* if detection were perfect. We assumed cases are detected with and independent and identical probability of *p*_*d*_ per case. Each primary case that was infected on day *i* as above was assigned a counterfactual detection date *k* at the same time its secondary cases were simulated. All cases were added to the count *d*_*k*_ of potential detections. For successful detections, no secondary cases whose infection date falls after day *k* were simulated. Time series generated by this process are assigned a likelihood score reflecting their similarity to the series of case counts reported by health departments. The count of reported new cases by day was smoothed for this purpose using a five day moving average. Given a series *C*_*i*_ of smoothed reported new cases by day, the likelihood was the binomial probability of detecting *C*_*i*_ cases given *d*_*i*_ cases to be detected and detection rate *p*_*d*_. The likelihood assigned to a simulation time series overall was constructed from the mean of daily log likelihoods over all the daily counts that were used in scoring. For scoring, case counts before the first date on which the number of new cases was greater than two were omitted, and all days from that day forward were used for scoring. The assignment of model days *i* to calendar dates was made by assigning the first day of simulation to a random date 1 to 21 days before the first scoring date, to allow for model populations with differing values of *R*_*t*_ to take differing amounts of time to arrive at case counts matching the scoring values.

### Epidemiological assumptions

We built our COVID model from an earlier simulation model, which was used to represent measles or Ebola virus disease transmission [42, 43]. In simulating transmission of SARS-CoV-2 from individual to individual, we used several assumptions.

We first assumed independent and identically distributed random times for the incubation interval, i.e., the number of days from infection of an individual to symptom onset. Given that interval, the time to case detection is an independent identically distributed offset from the date of symptom onset, as are all transmission events from that individual.

The number of secondary cases per case is assumed to be negative binomially distributed, and independent between cases. The mean number of cases per case (the effective reproduction number *R*_*t*_) is taken to be constant except for specific changes, reflecting recommendations for social distancing. Bay Area counties were recommended to begin social distancing on 17 March 2020. *R*_*t*_ takes a single value before 17 March and three constant values thereafter, in the same range, with discontinuous changes on 8 April and 18 April, representing the dates of adoption of contact tracing and cloth masks. The model selects values from those ranges based on their ability to reproduce the reported case counts in Bay Area counties.

Finally, the rate of detection of COVID-19 cases is unknown, and may be changing over time depending on test availability. Detection probability per individual ranges from 0.011 to 0.6 both before and after 10 March, and those two values are selected separately based on matching Bay Area case counts.

The model uses the above assumptions to generate stochastic time series of numbers of all infected individuals, and of the smaller numbers who are detected (reported). The detection rate is used as a binomial likelihood function to assign likelihoods to model timeseries and the parameter values that generated them, which is used in sampling/importance resampling [22] to construct a posterior distribution of parameters and outcomes. Simulation outcomes projected forward in time are used with this importance distribution to construct probabilistic forecasts.

### Parameterization

**The distribution of number of secondary cases per primary case** is assumed to be negative binomial, with mean *R*_*t*_ and dispersion parameter *k* [4]. The value of *R*_*t*_ can vary by day of transmission. We are assuming that *R*_*t*_ takes on four values, constant except for instantaneous changes on 17 March 2020, 8 April 2020, and 18 April 2020, reflecting the dates of introduction of San Francisco’s shelter in place policy, contact tracing program, and cloth mask policy, respectively. These constant values are uniformly distributed from 0.3 to 3.0 in each of these periods. The dispersion parameter *k* is assumed constant in time, at a value sampled uniformly from 0.1 to 0.8 (representing distributions with more tail density than the geometric distribution, as has been estimated in other diseases, i.e. presence of some frequency of superspreading events).

**The distribution of number of days from infection to symptom onset**, i.e. the incubation period, was assumed to be log-normally distributed with mean 5.6 days and standard deviation 4.2 days [16].

**The distribution of number of days from symptom onset to a transmission event**, which may be negative reflecting presymptomatic transmission, was modeled by −2.5 days plus a gamma distributed number of days with scale parameter 1.5 and shape parameter 2.1 [16]. This assumption was modified by using this distribution to model transmission events relative to either the symptom onset date or 3 days after infection, whichever is later. **The distribution of number of days from symptom onset to becoming a reported case**, the detection interval distribution, is assumed to be log-normally distributed with mean 8.9 days and standard deviation 7 days, to reflect the possibility that some cases may be detected soon after symptom onset and others may be detected several weeks later. **The detection probability** is assumed constant, taken from a uniform distribution from 0.011 to 0.6.

**One index case** is assumed to seed each simulation realization, with an infection date uniformly sampled 1 to 21 days before the first recorded case count used for likelihood scoring.

### Calculation of effective reproduction number

Central estimates and confidence intervals on daily counts and parameter values are constructed by importance sampling as follows. Likelihoods are transformed into a probability distribution by normalizing to 1:

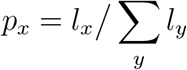

where *l*_*x*_ is the likelihood of realization *x*.

Estimates for future cumulative case counts are constructed using the importance sampled model series of new counts by day, by adding the cumulative new counts on days after the last reported date to the last reported count.

The effective reproduction number by day can be recorded in at least two ways. First, the effective reproduction number of each case with a given date of infection is the actual number of cases infected by that case,

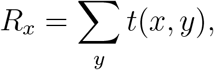

where *t*(*x, y*) is one if case *x* is the source of infection for case *y* infection, and zero otherwise. The effective “case reproduction number” on a given day is the average of those of the cases infected that day:

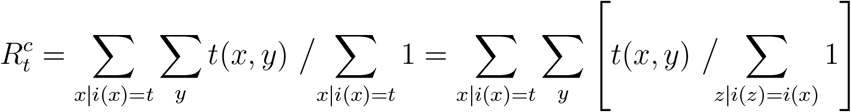

where *i*(*x*) is the date of infection of case *x*.

Second, a measure of *instantaneous* effective reproduction number by day [9], measuring transmission occurring on a given day rather than transmission from cases infected on a given day, but in units of cases per case, is

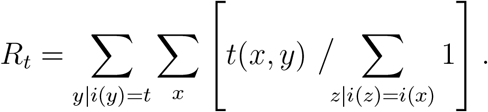

These two quantities averaged over all days of the model will give the same overall average *R*. The second estimator gives a more timely indication of changes in transmission rate. The model estimates of reproduction number by day reported in this paper are constructed using the latter method, computed from the actual number of transmission events generated by the simulation per day.

## Footnotes

1 https://www.sfdph.org/dph/alerts/files/HealthOrderC19-07-%20Shelter-in-Place.pdf, http://www.acphd.org/media/561969/faqs-order-shelter-in-place-20200324.pdf, retrieved May 3, 2020.

2 https://covid19.ca.gov/img/Executive-Order-N-33-20.pdf, retrieved May 3, 2020.

3 https://sfmayor.org/article/san-francisco-issues-new-policy-face-coverings, retrieved May 3, 2020.

4 https://github.com/nytimes/covid-19-data, last accessed April 30, 2020.

